# Change in vaccine willingness in Australia: August 2020 to January 2021

**DOI:** 10.1101/2021.02.17.21251957

**Authors:** Nicholas Biddle, Ben Edwards, Matthew Gray, Kate Sollis

## Abstract

The ANU Centre for Social Research and Methods ANU COVID-19 Impact Monitoring Survey Program asked the same group of respondents about their vaccine intentions in August 2020 and January 2021. The paper provides data on the vaccine willingness in Australia as of January 2021 and how this changed since August 2020 both at the national level and for particular individuals. The paper provides estimates of how vaccine willingness has changed for different population sub-groups and the individual level characteristics which are associated with changes in vaccine willingness. We find an overall decrease in vaccine willingness, with the biggest decline being those who would definitely get a vaccine as of August 2020 but said they would only probably get a vaccine as of January 2021. We also look at the factors associated with vaccine willingness, as well as the factors associated with change through time.

**Executive summary:**

- The paper provides data on the vaccine willingness in Australia as of January 2021 and how this changed since August 2020 both at the national level and for particular individuals.
- **There has been a substantial increase in vaccine resistance and hesitancy and a large decline in vaccine likeliness** between August 2020 and January 2021
  - Combined, 21.7 per cent of Australians said they probably or definitely would not get a safe and effective COVID-19 vaccine in January 2021, a significant and substantial increase from the 12.7 per cent of Australians who gave the same responses in August 2020.
- At the individual level, **31.9 per cent of Australians became less willing to get the vaccine between August 2020 and January 2021** in that they moved from a more to a less willing category.
  - There were still some Australians who became more willing over the period to get vaccinated (9.9 per cent).
- The largest single flow across willingness categories was the **18.7 per cent of Australians who went from being definitely willing to get a COVID-19 vaccination to only probably willing to get one**. There was a large decline in vaccine certainty, alongside increases in vaccine resistance.
- We found three attitudinal factors that were particularly important in explaining the decline in willingness. Those Australians who think too much is being made of COVID-19, those who have low confidence in hospitals and the health care system, and those who are not optimistic about the next 12 months had all decreased in terms of their willingness to get vaccinated once a vaccine is available.
  - In addition to campaigns targeting vaccination directly, **those programs that improve confidence, remind people of the dangers of COVID-19, but importantly highlight the potential for a much better 2022 all have the potential to improve vaccination rates**.
- Females, **Indigenous Australians, those who speak a language other than English at home and those who have not completed Year 12 have all became less willing** to get a vaccine since August 2020 compared to the rest of the Australian population.
  - These population groups are arguably the most urgent focus of any public health campaigns to improve willingness, both because they have low willingness to start with, but also because there is the potential opportunity to bring their willingness back to what it was in August 2020 when there was a smaller gap with the rest of the Australian population.
  - There is a real need to consider a significantly enhanced public health campaign in languages other than English
  - There is a need to convey information to the general public in a way that is informative, reassuring and salient for those without a degree

## 1 Introduction and overview

Australians, like much of the world, are hoping and waiting for a successful roll out of a vaccine for COVID-19 that would allow for a return to a more normal life. While Australia’s rate of infection and mortality is very low compared to many other countries, this has been achieved by severely limiting international travel including for Australian citizens wishing to return to Australia, and compulsory quarantine for arrivals from all parts of the world except for New Zealand. In addition, state and territory borders have been closed for periods of time and there have been “lock downs” in different parts of Australia in response to COVID outbreaks.

Significantly easing these restrictions will almost certainly require a sufficient number of Australians to have been vaccinated (in particular the most vulnerable) and international arrivals able to show they themselves are vaccinated or from a country with very low infection rates.

At the time of writing, in mid-February 2021, there were a number of vaccines that appear to have met the safe and effective threshold, albeit with some uncertainty around the effectiveness for some of the vaccine candidates for all population groups and for all of the variants of SARS-CoV-2 (Strizova 2021). A substantial number of countries had commenced vaccination with Israel having 71.6 COVID-19 doses per 100 population having been administered (people receive multiple doses so this is not the proportion of the population vaccinated), United Arab Emirates having 49.6 does per 100 population and the United Kingdom 21.4 doses per 100 population. ^1^

On the 25 January 2021 the Australian Therapeutic Goods Administration (TGA) provisionally approved the Pfizer/BioNTech COVID-19 vaccine for people aged 16 years of age and over. COVID-19 vaccination in Australia is scheduled to start in the second half of February 2021. In order for this vaccination program to be successful in achieving herd immunity a large enough proportion of the population need to be vaccinated. This makes monitoring the proportion of the population that intend to be vaccinated critical public health information. Perhaps even more importantly understanding how vaccination intentions differ between population sub-groups and the factors associated with changes in vaccine intentions requires a large dataset with individuals tracked through time.

The ANU Centre for Social Research and Methods ANU COVID-19 Impact Monitoring Survey Program asked the same group of respondents about their vaccine intentions in August 2020 and January 2021.2 In addition, the January 2021 survey asked a question on confidence in the vaccine development process. At the time of the survey in January 2021 a number of countries had started or were well into the process of vaccinating their populations using one or more of the recently developed vaccines. While the roll-out of the vaccines had not been completely smooth, there had at least not been any large-scale incidences of adverse effects from the vaccines currently being administered. While as noted above provisional approval for the use of the first vaccine in Australia was made on 25 January, there was a strong community expectation that approval was imminent.

The paper provides data on the vaccine willingness in Australia as of January 2021 and how this changed since August 2020 both at the national level and for particular individuals. The paper provides estimates of how vaccine willingness has changed for different population sub-groups and the individual level characteristics which are associated with changes in vaccine willingness.

The remainder of the paper is structured as follows. In the next section, we summarise the international data on vaccine willingness/hesitancy in January 2021. In Section 3 we look at changes at the national level and in Section 4 we look at the individual trajectories for vaccine willingness between August 2020 and January 2021. Section 4 looks in detail at the factors that are associated with that change. Section 5 concludes.

## 2 International evidence on vaccine hesitancy

Internationally, preliminary evidence suggests that while vaccine hesitancy initially decreased in the early stages of the pandemic (Caserotti et al. 2021), it has been on the rise in a number of countries throughout the world, even those experiencing high rates of COVID-19, and those under lock-down conditions. For example, Wang et al. (2021) found that vaccine acceptance in Hong Kong was lower during their third COVID-19 wave, compared to the first, and research by Hacquin et al. (2020) suggests that vaccine refusal steadily rose between May and September in France, with only 23% of survey participants being willing to take a future vaccine as reported in September 2020. Similarly, findings from the United States indicate that there was a steady increase in vaccine hesitancy throughout 2020 (Firdman et al. 2020; Huynh 2020; Daly & Robinson 2020), however recent research by Pew Research Center (2020) suggests that this trend might be starting to change, with 60% of Americans reporting that would get a vaccine for COVID-19 in November 2020, up from 51% in September.

The fact that vaccine hesitancy appears to be rising in countries heavily affected by COVID-19 contradicts previous research which suggests that the increased risk of a disease should result in improvements to vaccine acceptance (Fridman et al. 2020). Potential explanations for a rising trend in vaccine hesitancy include greater safety concerns and a perceived rush of vaccine development (Daly et al. 2020; Huynh 2020; Wang et al. 2021), a lack of trust in government and pharmaceutical industries, and perceptions that the vaccine isn’t effective (Hacquin et al. 2020).

## 3 Vaccine willingness and confidence

The August 2020 ANUpoll included the question ‘If a safe and effective vaccine for COVID-19 is developed, would you…definitely not get the vaccine, probably not get the vaccine, probably get the vaccine, or definitely get the vaccine? The January 2021 survey included a very similar question ‘When health officials notify the public that a safe and effective COVID-19 vaccine is available in Australia, will you…?’, with the same response options as of August 2020.^3^

We find a significant increase in vaccine hesitancy. In August 2020, 5.5 per cent of adult Australians said that they would definitely not get the vaccine, 7.2 per cent said that they would probably not get the vaccine, 28.7 per cent said that they would probably get the vaccine and 58.5 per cent said that they would definitely get the vaccine (Edwards et al. 2020).

In January 2021, 8.4 per cent of Australians said they would definitely not get the vaccine and 13.3 per cent said they probably would not get a vaccine once health officials notify that a vaccine is available. This is a substantial increase from the same response options in August 2020. There was also an increase in the per cent of Australians who said they would only ‘probably’get the vaccine, from 28.7 per cent in August 2021 to 34.7 per cent during the most recent survey. The only option that has declined, therefore, is those who said they would ‘definitely’get the vaccine, down from 58.5 per cent in August 2020 to 43.7 per cent in January 2021.

There has been a substantial increase in vaccine resistance and hesitancy and a large decline in vaccine likeliness between the time when a vaccine was still being developed and January 2021 when a number of vaccines were available and being administered internationally, but not yet available in Australia.

In addition to asking people about their own willingness to take a COVID-19 vaccination once one is available, we also asked respondents in January 2021 ‘How much confidence, if any, do you have that the research and development process will produce a vaccine for COVID-19 that is safe and effective?’. Across the adult Australian population, we estimate that around a quarter of Australians (25.2 per cent) have ‘A great deal of confidence’with about half of the population (49.6 per cent) having ‘A fair amount of confidence.’The remaining quarter of the population can be broken down into the 18.5 per cent who have ‘Not too much confidence’and 6.7 per cent who have ‘No confidence at all.’

Perhaps not surprisingly there is a strong correlation between vaccine willingness and confidence in the research and development process with the higher the level of confidence in the research and development process the greater higher the level of vaccine willingness.4 Of those who had a great deal or a fair amount of confidence in the research and development process, 82.0 per cent said they definitely or probably would get vaccinated. For those who have not too much or no confidence at all, on the other hand, this declines to 31.9 per cent.

## 4 Changes in vaccine willingness at the individual level

As reported above, there has been an increase in vaccine resistance and vaccine hesitancy in Australia since August 2020. One of the strengths of the data used in this paper is that its longitudinal nature allows change to be analysed at the individual respondent level. Specifically, there are 2,737 respondents who completed the vaccine question in both August 2020 and January 2021.^5^

Figure 1 gives the flows between a person’s vaccination intentions when asked in August 2020 and in January 2021 using those who responded to both surveys. The numbers after the category names give the (weighted) per cent of the longitudinal sample in each of the categories at that particular point in time, with the size of the individual ‘ribbons’proportional to the sizes of the flows.

**Figure 1.**
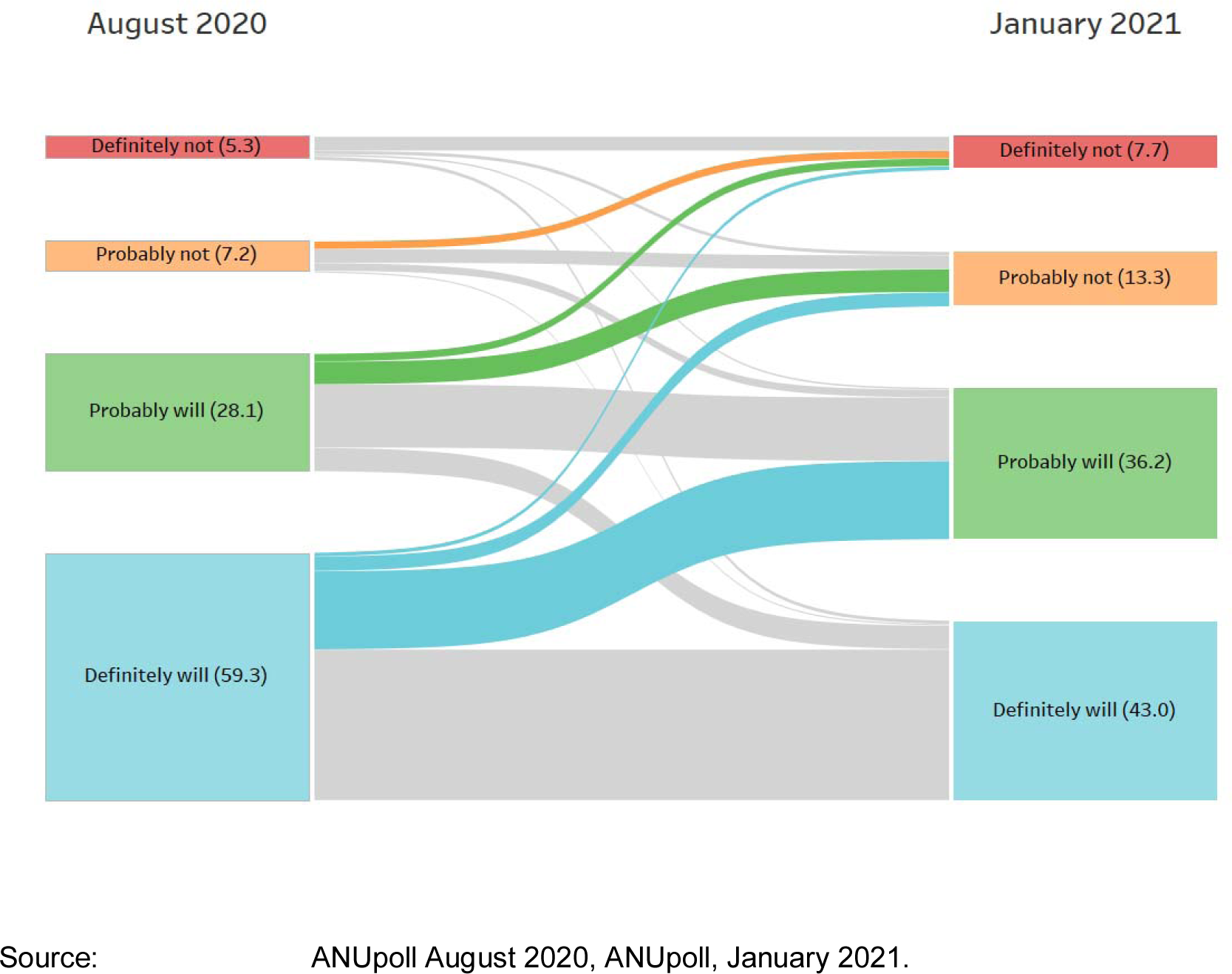
Changes in vaccine willingness between August 2020 and January 2021.

There are several points to take from Figure 1. First, confirming the repeated cross-sectional results presented earlier, there is a greater flow from more to less willing (the coloured ribbons moving upwards in the diagram) than there is movement from less to more willing. At the individual level, 31.9 per cent of Australians became less willing to get the vaccine between August 2020 and January 2021 in that they moved from a more to a less willing category. The second thing to note from the diagram is that there were still some Australians who became more willing over the period to get vaccinated (9.9 per cent), it is just that this group was far outweighed by those who moved in the opposite direction.

The final key point to note from the diagram is the largest single flow was the 18.7 per cent of Australians who we estimate went from being definitely willing to get a COVID-19 vaccination to only probably willing to get one. Not only is this the largest change in absolute terms, but it is also the largest change as a per cent of the baseline population. Put simply, one of the biggest concerns from a herd immunity perspective between mid 2020 and early 2021 is the very large increase in vaccine uncertainty.

## 5 Explaining vaccine willingness and changes

### 5.1 Vaccine willingness in January 2021

One of the key policy challenges with regards to the roll-out of vaccines is that vaccine willingness rates are quite different across the country. However, this also creates an opportunity, in that resources and messaging campaigns can be targeted towards those groups with lower rates of acceptance.

In order to better understand the demographic, socioeconomic, and geographic factors associated with vaccine willingness and confidence in the research and development process regression models have been estimated. These models allow the effects of individual variables to be estimated while holding constant the effects of other variables.

Given that the dependent variables (vaccine willingness and confidence in vaccine research and development processes) take one of four mutually-exclusive values each and these have a natural ordering, an ordered probit regression model is used (Table 1). In these models, positive values indicate a greater willingness or confidence than the base case which is described in the note to Table 1 and negative values indicate a lower willingness or confidence. The direction and the magnitude of the coefficients are very similar for the willingness regression and the confidence regression. The factors that are associated with a person’s willingness to get a vaccine (at least as revealed in a social survey) are very similar to the factors associated with a person’s confidence in the research and development process for developing a safe and effective vaccine. We therefore focus on the willingness variable in the discussion of the model estimates.

**Table 1.**
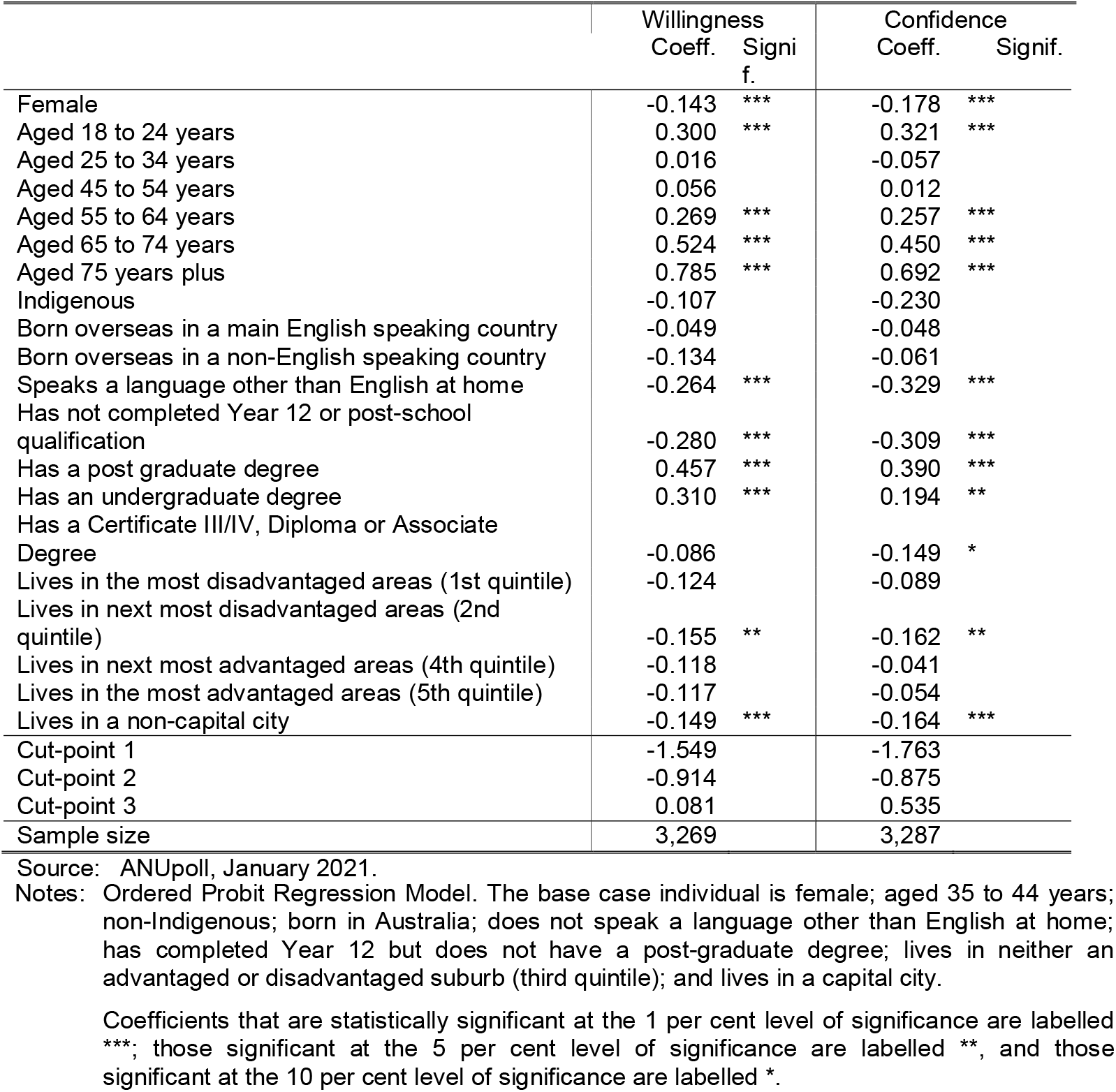
Demographic, socioeconomic, and geographic factors associated with vaccine willingness and confidence in the research and development process

|  | Willingness<br>Coeff. Signi<br>f. | Confidence<br>Coeff. Signif. |
| --- | --- | --- |
| Female | -0.143 *** | -0.178 *** |
| Aged 18 to 24 years | 0.300 *** | 0.321 *** |
| Aged 25 to 34 years | 0.016 | -0.057 |
| Aged 45 to 54 years | 0.056 | 0.012 |
| Aged 55 to 64 years | 0.269 *** | 0.257 *** |
| Aged 65 to 74 years | 0.524 *** | 0.450 *** |
| Aged 75 years plus | 0.785 *** | 0.692 *** |
| Indigenous | -0.107 | -0.230 |
| Born overseas in a main English speaking country | -0.049 | -0.048 |
| Born overseas in a non-English speaking country | -0.134 | -0.061 |
| Speaks a language other than English at home | -0.264 *** | -0.329 *** |
| Has not completed Year 12 or post-school qualification | -0.280 *** | -0.309 *** |
| Has a post graduate degree | 0.457 *** | 0.390 *** |
| Has an undergraduate degree | 0.310 *** | 0.194 ** |
| Has a Certificate III/IV, Diploma or Associate Degree | -0.086 | -0.149 * |
| Lives in the most disadvantaged areas (1st quintile) | -0.124 | -0.089 |
| Lives in next most disadvantaged areas (2nd quintile) | -0.155 ** | -0.162 ** |
| Lives in next most advantaged areas (4th quintile) | -0.118 | -0.041 |
| Lives in the most advantaged areas (5th quintile) | -0.117 | -0.054 |
| Lives in a non-capital city | -0.149 *** | -0.164 *** |
| Cut-point 1 | -1.549 | -1.763 |
| Cut-point 2 | -0.914 | -0.875 |
| Cut-point 3 | 0.081 | 0.535 |
| Sample size | 3,269 | 3,287 |

Source: ANUpoll, January 2021.
Notes: Ordered Probit Regression Model. The base case individual is female; aged 35 to 44 years; non-Indigenous; born in Australia; does not speak a language other than English at home; has completed Year 12 but does not have a post-graduate degree; lives in neither an advantaged or disadvantaged suburb (third quintile); and lives in a capital city.
Coefficients that are statistically significant at the 1 per cent level of significance are labelled \*\*\*; those significant at the 5 per cent level of significance are labelled \*\*, and those significant at the 10 per cent level of significance are labelled \*.

Consistent with other literature, females are less willing to get a vaccine than males. This is demonstrated by the negative coefficient in the more detailed econometric modelling in Table 1, but is also evident in the simple descriptive statistics (Figure 2). Without controlling for other characteristics, we estimated that only 39.9 per cent of females think they will definitely get a safe and effective vaccine, compared to 47.4 per cent of males. There is a larger percentage of females who think they will probably get the vaccine (36.8 per cent) than males (32.6 per cent), but even when combined there are fewer females than males in proportionate terms who feel they are more likely than not to get the vaccine.

**Figure 2.**
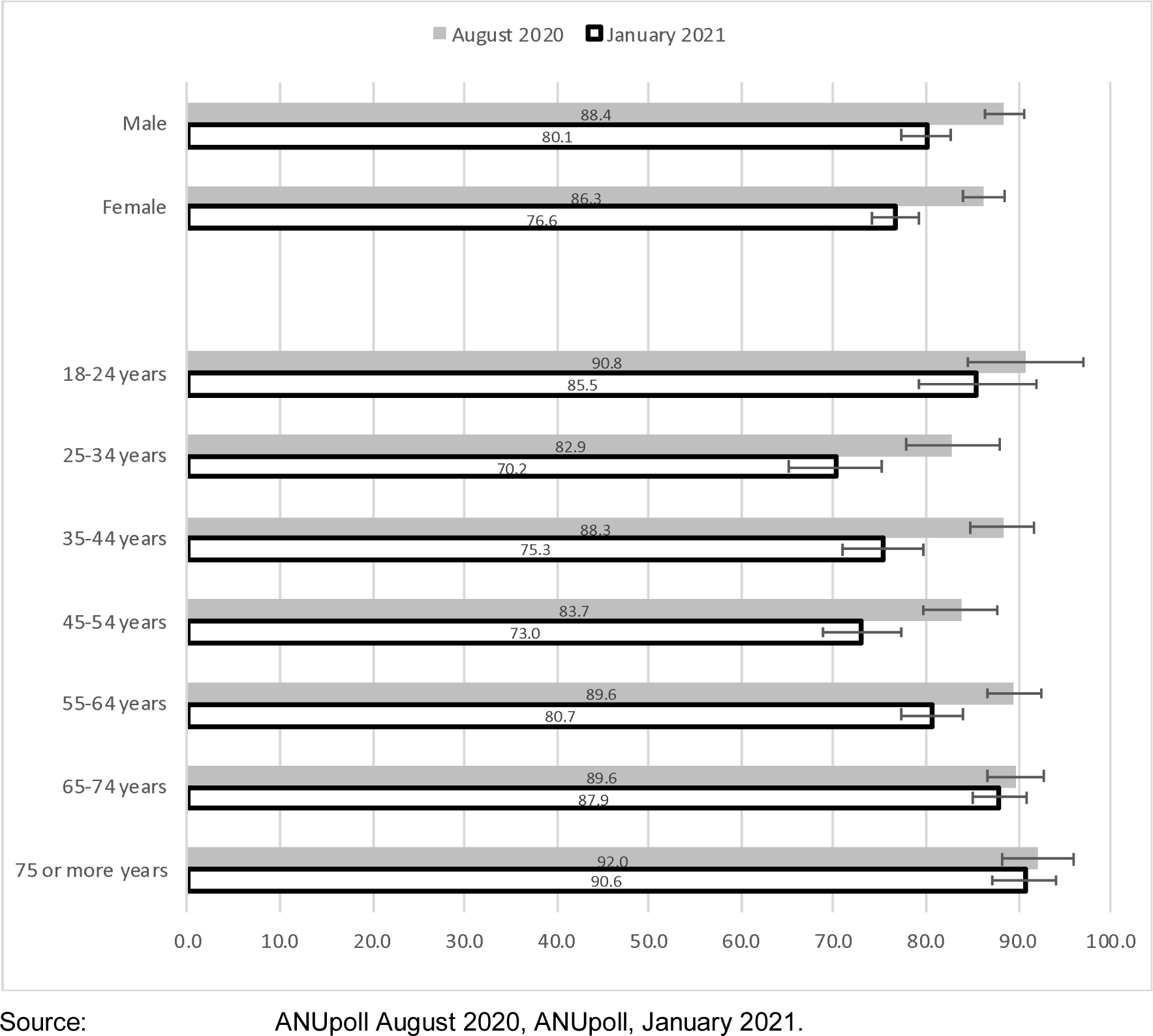
Vaccine willingness between August 2020 and January 2021, by sex and age.

There are also significant differences by age. Young Australians (aged 18 to 24 years) and older Australians (aged 55 years and over) have the highest levels of willingness. To put this gap in perspective, 85.5 per cent of 18-24 year olds and 90.6 per cent of those aged 75 years and over say they definitely or probably will get vaccinated. Only 70.2 per cent of those aged 25 to 34 years say they will.

Of the other demographic variables, there is no significant difference between Indigenous and non-Indigenous Australians (although there is a small negative coefficient, that is not statistically significant). This is a promising finding that, although needing to be validated with more targeted datasets, does give some hope that a relatively vulnerable population sub-group that may be more susceptible to COVID-19 if infected is at least as willing to get vaccinated as the general population. There are also no major differences by migration status.

One of the more concerning findings from a public health perspective is that those who speak a language other than English at home are significantly less willing to get vaccinated than those who speak English only. Given our surveys are only conducted in English, we are likely to be missing those with the lowest levels of English language ability. Nonetheless, it does highlight a real need to target messages to this group and to consider a significantly enhanced public health campaign in languages other than English.

We find that education levels have a significant and substantial association with vaccine willingness. Year 12 completion does have a significant association (compared to those who have completed Year 12, all else being equal), but it is degree qualifications that are the most predictive of vaccine willingness. Without controlling for other characteristics, 86.9 per cent of those with a postgraduate degree say they will probably or definitely get vaccinated, alongside 84.6 per cent of those with a bachelor’s degree only. For those without a degree, however, this declines to 75.6 per cent, highlighting the need to convey information to the general public in a way that is informative, reassuring and salient for those without a degree.

There is a lower levels of vaccine willingness for those who live outside capital cities compared to those who live within a capital city. The difference in raw percentages is not as large as it is for some of the other variables, implying that some of the differences in the econometric model are due to observed characteristics. Nonetheless, in the econometric model there is a negative and significant coefficient, and when making bivariate comparisons, only 79.4 per cent of those in a capital city say they probably or definitely will get the vaccine compared to 76.1 per cent of those in a non-capital city. This may be due in part to a consistently lower level of infections in less urban parts of the country. Nonetheless, infection risk will still remain for non-capital cities and regional and remote areas in general, and may even increase as vaccinated Australians from capital cities and urban areas travel around the country without having been tested and without symptoms. Once again, a targeted set of information for these geographic areas is worth considering from a policy perspective.

### 5.2 Explaining changes in vaccine willingness

The analysis in the previous section is of the demographic, socioeconomic, and geographic factors associated with vaccine willingness and confidence in January 2021. Some, but not all of the above associations are similar to those found in previous waves of ANUpoll (Edwards et al. 2020) and are therefore picking up stable characteristics associated with vaccine willingness (albeit with different magnitudes).

This section extends this analysis to look at the characteristics of the population that predict changes through time in vaccine willingness and, in particular, the decrease in vaccine willingness observed between August 2020 and January 2021. We follow a number of complimentary approaches to understand what factors are associated with changes in vaccine willingness.

The first step is to estimate a regression model with the dependent variable being vaccine willingness in January 2021 including all of the demographic, geographic and socio-economic characteristics included in the models reported in Table 1 plus a range of other variables related to attitudes, beliefs and circumstances. We then progressively remove variables or sets of variables that have the lowest level of statistical significance until all explanatory variables are significant at least at the 10 per cent level of significance.^6^

With this set of explanatory variables, four complimentary econometric models are run. The first model is for vaccine willingness including all four categories (i.e., same as in the model reported in Table 1), but including the broader set of explanatory variables. The second model adds to the first model vaccine willingness in August 2020 and is thus an estimate of the factors associated with changes in vaccine willingness between August 2020 and January 2021 (i.e., a lagged dependent variable model).

The third model is of the factors associated with becoming less willing between August 2020 and January 2021. This model excludes those who said, in August 2020 that they definitely would not get a vaccine. This model compares those who are become less vaccine willing (i.e., the coloured ribbons in Figure 1 or 33.7 per cent of the relevant in scope population) to those whose vaccine willingness did not change or they become more vaccine willing. The fourth model looks at those who said that they would definitely or probably get the vaccine in August 2020 and models the factors associated with saying they would probably or definitely **not** get the vaccine in January 2021. That is, those who flipped from being on balance willing to on balance not willing.

#### 5.2.1 Vaccine willingness (January 2021 ANU Poll Survey only)

Beginning with the results presented in the first column (the expanded cross-sectional analysis) there are a range of more time varying attitudinal and outcome variables that are associated with vaccine willingness. Those who had been tested for COVID-19 were more likely to say they would get vaccinated, as were those who had experienced anxiety or worry due to COVID-19. This highlights the role that a person’s own COVID-related experience plays in their vaccine willingness. There is a very strong negative relationship between thinking too much is being made of COVID-19 and vaccine willingness, highlighting the need to remind people (in a careful and targeted way) of the ongoing potential effects of COVID-19, even if Australia has been relatively successful in managing infections and mortality.

There was a positive relationship between confidence in the Federal government and an even stronger association with confidence in State/Territory governments, who are more responsible for public health delivery in Australia. The institution that has the strongest association though is hospitals and the health care system. To put these results in perspective, 80.9 per cent of those Australians who say they are confident in hospitals and the health system say they probably or definitely will get vaccinated, compared to only 63.0 per cent of those who are not confident.

Those who have experienced financial stress in the last 3 months (as indicated by not being able to pay their rent or mortgage on time) are much less likely to say they will get vaccinated. Clearly, people in those circumstances have very pressing immediate concerns, and the short-term risk of illness from the vaccine may be more salient whereas the medium to long-term benefit of the vaccine may be of less relevance. Finally, those who are more optimistic about the future or who have a belief in a strong role of government are more likely to say they will get vaccinated.

#### 5.2.2 Vaccine willingness – changes from August 2020

In the second column of results in Table 2 we add vaccine intentions from August 2020 so that now the variables in the model show how these variables are associated with changes through time in vaccination willingness. Fewer variables were statistically significant with being tested for COVID-19, being anxious or worried due to COVID-19, confidence in Federal or state or territory governments and being born overseas in a non-English speaking country were not associated with changes in vaccine intentions. In part, this may be due to the smaller sample size (due to the use of the longitudinal sample) but also suggests that some variables predict stable traits, rather than explain short-term changes in intentions.

**Table 2.** Factors associated with vaccine willingness and change in vaccine willingness, August 2020 and January 2021

|  | Willingness<br>(categorical I)<br>Coeff. Signif. |  | Willingness<br>(categorical II)<br>Coeff. Signif. |  | Decreased<br>willingness<br>Coeff. Signif. | No longer willing<br>Coeff. Signif. |
| --- | --- | --- | --- | --- | --- | --- |
| Definitely not get vaccine as of August 2020 |  |  | -2.037 *** |  |  |  |
| Probably not get vaccine as of August 2020 |  |  | -1.737 *** |  | -0.534 *** |  |
| Probably get vaccine as of August 2020 |  |  | -0.898 *** |  | -0.481 *** | 0.748 *** |
| Tested for COVID-19 | 0.121 ** |  | 0.089 |  | -0.081 | -0.156 |
| Anxious or worried due to COVID-19 | 0.234 *** |  | 0.056 |  | -0.017 | -0.005 |
| Thinks too much is being made of COVID-19 | -0.451 *** |  | -0.293 *** |  | 0.222 ** | 0.087 |
| Confident in the Federal government | 0.110 * |  | 0.062 |  | -0.042 | -0.199 * |
| Confident in State/Territory government | 0.159 ** |  | 0.073 |  | -0.093 | -0.017 |
| Confident in hospitals and health care system | 0.286 *** |  | 0.231 *** |  | -0.207 ** | -0.241 * |
| Unable to pay rent or mortgage in previous 3 months | -0.256 *** |  | -0.255 *** |  | 0.125 | 0.261 * |
| Thinks life will be improved in next 12 months | 0.195 *** |  | 0.203 *** |  | -0.230 *** | -0.225 ** |
| Index of belief in strong role of government | 0.025 *** |  | 0.018 ** |  | -0.023 ** | -0.021 |

Change in vaccine willingness – August 2020 to January 2021
|  | Willingness<br>(categorical I) |  | Willingness<br>(categorical II) |  | Decreased<br>willingness |  | No longer willing |  |
| --- | --- | --- | --- | --- | --- | --- | --- | --- |
|  | Coeffic. | Signif. | Coeffic. | Signif. | Coeffic. | Signif. | Coeffic. | Signif. |
| Female | -0.258 | *** | -0.217 | *** | 0.254 | *** | 0.012 |  |
| Aged 18 to 24 years | 0.270 | ** | 0.207 |  | -0.026 |  | 0.064 |  |
| Aged 25 to 34 years | -0.051 |  | -0.074 |  | 0.101 |  | 0.354 | ** |
| Aged 45 to 54 years | 0.036 |  | 0.025 |  | -0.046 |  | 0.104 |  |
| Aged 55 to 64 years | 0.306 | *** | 0.180 | ** | -0.153 |  | -0.051 |  |
| Aged 65 to 74 years | 0.510 | *** | 0.329 | *** | -0.290 | ** | -0.309 | * |
| Aged 75 years plus | 0.794 | *** | 0.533 | *** | -0.529 | *** | -0.463 | ** |
| Indigenous | -0.128 |  | -0.270 |  | 0.452 | ** | 0.365 |  |
| Born overseas in a main English speaking country | -0.077 |  | 0.047 |  | -0.078 |  | 0.049 |  |
| Born overseas in a non-English speaking country | -0.168 | ** | -0.145 |  | 0.129 |  | 0.252 |  |
| Speaks a language other than English at home | -0.239 | *** | -0.213 | ** | 0.255 | ** | 0.005 |  |
| Has not completed Year 12 or post-school qualification | -0.152 |  | -0.235 | ** | 0.254 | ** | 0.215 |  |
| Has a post graduate degree | 0.431 | *** | 0.255 | ** | -0.163 |  | -0.215 |  |
| Has an undergraduate degree | 0.288 | *** | 0.119 |  | -0.089 |  | -0.123 |  |
| Has a Certificate III/IV, Diploma or Associate Degree | -0.044 |  | -0.161 | * | 0.183 |  | 0.264 | * |
| Lives in the most disadvantaged areas (1st quintile) | -0.095 |  | 0.062 |  | -0.134 |  | 0.012 |  |
| Lives in next most disadvantaged areas (2nd quintile) | -0.147 | * | -0.079 |  | -0.019 |  | 0.126 |  |
| Lives in next most advantaged areas (4th quintile) | -0.118 |  | 0.139 |  | -0.128 |  | -0.152 |  |
| Lives in the most advantaged areas (5th quintile) | -0.139 | * | -0.061 |  | -0.014 |  | 0.124 |  |
| Lives in a non-capital city | -0.126 | ** | -0.100 |  | 0.066 |  | 0.081 |  |
| Cut-point 1 | -0.996 |  | -2.083 |  |  |  |  |  |
| Cut-point 2 | -0.300 |  | -1.220 |  |  |  |  |  |
| Cut-point 3 | 0.756 |  | 0.070 |  |  |  |  |  |
| Constant |  |  |  |  | 0.054 |  | -1.167 |  |
| Sample size | 3,184 |  | 2,562 |  | 2,433 |  | 2,279 |  |

Source: ANUpoll, August 2020 and January 2021.
Notes: Probit Regression Model. The base case individual is female; aged 35 to 44 years; non-Indigenous; born in Australia; does not speak a language other than English at home; has completed Year 12 but does not have a post-graduate degree; lives in neither an advantaged or disadvantaged suburb (third quintile); and lives in a capital city. Coefficients that are statistically significant at the 1 per cent level of significance are labelled \*\*\*, those significant at the 5 per cent level of significance are labelled \*\*, and those significant at the 10 per cent level of significance are labelled \*.

For those that remained statistically significant, some coefficient values also declined between Model 1 and Model 2 (the lagged dependent variable model) but were still robustly associated with changes in intentions since August 2020. These included thinks too much is being made about COVID-19, confidence in hospital and health care system, financial stress (i.e. paying rent or mortgage on time), optimistic that life will improve in the next 12 months, belief in strong government, age and education.

#### 5.2.3 Decrease in willingness (excluding those who definitely would not have got the vaccine in August 2020)

Focusing on decreases in vaccine willingness (the third model), a few select variables were statistically significant, even after controlling for levels of willingness in August 2020. Those who think too much is being made of COVID-19 have become less willing through time (perhaps a reflection of the relative success of managing infections since August 2020), whereas those who have confidence in the hospitals and health care system have become more willing. Optimism about life in 2022 and a belief in a strong role of government were also associated with a higher probability of becoming willing to get vaccinated since August 2020.

Of the more stable explanatory variables, females, Indigenous Australians, those who speak a language other than English at home and those who have not completed Year 12 have all became less willing to get a vaccine since August 2020.

#### 5.2.4 No longer willing (changed from definitely or probably in August 2020 to probably or definitely not in January 2021)

Those that switched their vaccine intentions were more likely to be unable to pay their mortgage or rent in the last 3 months, more likely to be 45-54 years (compared to 35 to 44 years) and had a Certificate III/IV, Diploma or Associate Degree. Those that shifted to vaccine hesitancy (probably or definitely not in January 2021) were less likely to be:

- confident in the Federal Government;
- confident in hospitals or health care system;
- optimistic about the next 12 months; and
- 65 or more years.

## 6 Concluding comments

Australia has been ranked by the Lowy Institute as the 8^th^ most effective country (out of 98) in terms of COVID-19 policy response, based on confirmed deaths, cases and tests.^7^ While there have no doubt been large economic and social costs from that public health response, data published previously from ANUpoll has shown that life satisfaction and wellbeing in Australia is now back to levels observed prior to the spread of COVID-19, and that economic outcomes are close to having converged.

That does not, of course, mean that life in Australia is back to a pre-COVID normality, or that it will be any time soon. For this to occur, a much higher degree of herd immunity is likely to be necessary, and this is only likely to occur following an effective vaccination roll-out, reaching the vast majority of Australians, and the most vulnerable Australians in particular. Australia has been for the most part a bystander in the vaccine development process, but will have to have an effective public health response to ensure the faster than expected development of vaccinations translates into positive outcomes in Australia.

Data presented in this paper has shown that this may be becoming increasingly more challenging than first thought, as rates of vaccine willingness appear to be declining, with a particularly large decrease in vaccine certainty. Moreover, other research suggests that estimates of the time taken to the vaccinate the population may be too optimistic (Hanly et al., 2021), enabling more time for misinformation which in turn could drive vaccine hesitancy.

We found three attitudinal factors that were particularly important in explaining the decline in willingness. Those Australians who think too much is being made of COVID-19, those who have low confidence in hospitals and the health care system, and those who are not optimistic about the next 12 months had all decreased in terms of their willingness to get vaccinated once a vaccine is available. In addition to campaigns targeting vaccination directly, those programs that improve confidence, remind people of the dangers of COVID-19, but importantly highlight the potential for a much better 2022 all have the potential to improve vaccination rates.

In addition, females, Indigenous Australians, those who speak a language other than English at home and those who have not completed Year 12 have all became less willing to get a vaccine since August 2020. These population groups are arguably the most urgent focus of any public health campaigns to improve willingness, both because they have low willingness to start with, but also because there is the potential opportunity to bring their willingness back to what it was in August 2020 when there was a smaller gap with the rest of the Australian population.

## Data Availability

Data will be available through the Australian Data Archive.

doi:
10.26193/HQX1FV

## Acknowledgements

The authors would like to thank a number of people who were involved in the development of the ANUpoll questionnaires, including Diane Herz, Dr Benjamin Phillips, Dr Paul Myers, Matilda Page, Diana Nguyen, Anna Lethborg and Charles Dove from the Social Research Centre, and Professor Ian McAllister from the ANU. Financial support for the ANU COVID-19 Impact Monitoring Survey Program has been provided by the Australian Institute of Health and Welfare. For this particular survey, the ANU received funding from the Minderoo Foundation, for which we are particularly appreciative. We would also like to thank Dinith Marasinghe for assistance with tables and figures.

## Appendix 1 About the survey

The primary source of data for this paper is the January ANUpoll. The Social Research Centre collected data online and through Computer Assisted Telephone Interviewing (CATI) in order to ensure representation from the offline Australian population. Around 4.9 per cent of interviews were collected via CATI. The contact methodology adopted for the online Life in Australia^™^ members is an initial survey invitation via email and SMS (where available), followed by multiple email reminders and a reminder SMS. Telephone non-response of panel members who have not yet completed the survey commenced in the second week of fieldwork and consisted of reminder calls encouraging completion of the online survey.

The contact methodology for offline Life in Australia^™^ members was an initial SMS (where available), followed by an extended call-cycle over a two-week period. A reminder SMS was also sent in the second week of fieldwork.

A total of 4,055 respondents were invited to take part in the survey, leading to a wave-specific completion rate of 85.3 per cent. Taking into account recruitment to the panel, the cumulative response rate for this survey is around 7.3 per cent.

Unless otherwise stated, data in the paper is weighted to population benchmarks. For Life in Australia^™^, the approach for deriving weights generally consists of the following steps:

1. Compute a base weight for each respondent as the product of two weights:
  a. Their enrolment weight, accounting for the initial chances of selection and subsequent post-stratification to key demographic benchmarks
  b. Their response propensity weight, estimated from enrolment information available for both respondents and non-respondents to the present wave.
2. Adjust the base weights so that they satisfy the latest population benchmarks for several demographic characteristics.

The ethical aspects of this research have been approved by the ANU Human Research Ethics Committee (2014/241).

The previous waves of data collection consisted of a 15-20 minute survey, with the October 2020 survey slightly less than five minutes in length. A full-length survey was conducted in November 2020 with a further survey scheduled for April 2021.

A high proportion of respondents to the January survey (85.9 per cent) had been interviewed at least once since January 2020, with a number of new participants added to replace those who dropped out of Life in Australia™ over time and thus to maintain its representativeness or the Australian population. A slightly lower proportion of the sample (80.9 per cent) were interviewed in the November 2020 sample specifically, meaning we have a very large sample of Australians for whom we can track outcomes over the COVID-19 period, as well as over the two months preceding the survey.

Between the 18^th^ of January and the 1^st^ of February 2021, the Social Research Centre on behalf of the ANU Centre for Social Research and Methods undertook an ANUpoll as part of the sixth wave of the ANU’s COVID-19 Impact Monitoring Survey Program.

## Endnotes

1. Data is for up to 12th February and is from https://ourworldindata.org/covid-vaccinations
2. These surveys are being undertaken on a representative sample of using the Life in AustraliaTM nationally representative online panel which is collecting data from the same group of Australians. Surveys had been conducted with the same group of respondents (apart from the refresh members) in January and February 2020, just before the COVID-19 pandemic started in Australia and in April, May, August (when the last vaccination question was asked), October, and November after the pandemic started to cause major impacts in Australia, as well as during and just after the second wave of infections that were concentrated on Victoria. Full details of the survey are given in Appendix 1, with the survey itself soon to be available through the Australian Data Archive.
3. The change in question wording was required given that by January 2021 COVID-19 vaccinations were taking place in a number of countries due to the assessment of these countries that a safe and effective vaccination is available.
4. Across the four categories of each, the polychoric correlation is 0.812.
5. Between November 2020 and January 2021 the panel was refreshed with n = 385 panellists being retired and n = 612 new panellists being recruited. The longitudinal sample comprises 89.7 per cent of the 3,052 individuals that were interviewed in August 2020 and for whom we have vaccine data. Given this it is appropriate to use the August 2020 weights for the longitudinal analysis.
6. In order, we tested and then removed the following variables from the model:

- Serious or moderate levels of psychological distress, as measured by the K-6 index and thresholds (Kessler et al. 2002);
- A person’s own expected likelihood of being infected in the next 6 months;
- Number of hours worked per week in January 2021; and
- Confidence in the public service.

7 https://interactives.lowyinstitute.org/features/covid-performance/

